# Prevalence of Malaria from Blood Smears Examination: A Three-Year Retrospective Study from Nakfa Hospital, Eritrea

**DOI:** 10.1101/2020.03.24.20042747

**Authors:** Yafet Kesete, Shewit Mhretab, Meron Tesfay

**Affiliations:** Nakfa Hospital, Nakfa, Eritrea; Dep. of Clinical Laboratory Science, Asmara College of Health Sciences, Asmara, Eritrea

## Abstract

**Background:** Malaria afflicts more than 90 countries in the tropical and subtropical region in which more than half of cases are present in sub-Saharan, Africa. It is one of the major health issues in Eritrea in which about 3.6 million (71%) of Eritrean population live in areas conducive for high transmission of malaria. Studying its prevalence is necessary to implement effective control measures. Therefore, this study was conducted to determine the three-year prevalence of malaria from peripheral blood smear examination.

**Methods:** A retrospective study was conducted at Nakfa Hospital from September 2016 to August 2019. All demographic details of subjects and positive malaria cases data were collected from laboratory registration book.

**Results:** The overall prevalence of smear positive malaria cases was 33.0% (1921 cases out of 5826). Males (58.19%) were more prone to have a positive malaria smear than females (41.8%). A higher prevalence of malaria was observed in the adult age group (35%) compared with children under 5 years old children (27%). The highest prevalence of malaria cases was found in the year 2016 in which correspond to 48.8%. A comparative incidence was also observed during the next year of 2017 with rate of 41% (1087 cases out of 2680) and decreased to low levels in recent couple of years. High slide positive rate was seen in summer (40%) and Autumn (39.52%) whereas Spring had the lowest frequency of cases (14.3%). Similarly, months of July (45%) and September (40.6%) had been noted to have the maximum number of cases. *Plasmodium vivax* constituted the most predominant malaria infections (78.06%), while markedly lower *p. falciparum* was also present (21.26 %). Almost around 70% of cases were reported from patients living in and at the peripheries of Nakfa town.

**Conclusion:** Eritrea is one of the few countries which has made a profound progress on decreasing transmission of common communicable diseases. However, vigilant surveillance is required especially during malaria transmission peaks from July to October which also overlap with harvesting seasons in Eritrea. Therefore, health planners need to organize intensive health education to increase community awareness via promotion of information and communication.

## Introduction

During 19^th^ & 20^th^ century, Malaria, an endemic disease, was successfully eradicated in Europe and North America. Nevertheless, this has not been duplicated anywhere in tropical countries and especially in sub-Saharan countries of Africa. Malaria afflicts more than 90 countries in the tropical and subtropical region in which more than half of cases are present in sub-Saharan, Africa [1]. Moreover, about one third of the world is prone to infections caused by malarial species [2]. Estimates of World Health Organization indicate 300-500 million malaria cases occur yearly, with almost all of cases burden being in Africa. Similarly, from 700,000 to 2.7 million people die because of malaria yearly and >75% of them being African children and pregnant women [3].

Nowadays, approximately about 10% of the world population is a reservoir for malaria parasites in their bloodstream [4]. An estimated 219 million cases of malaria occurred worldwide in 2017 compared with 239 million cases in 2010 and 217 million cases in 2016 [3]. The African Region still bears the largest burden of malaria morbidity, with 200 million cases (92%) in 2017. Almost 80% of all malaria cases globally occur in 15 African countries and in India [3].

Malaria is one of the major health issues in Eritrea. The various eco-climatic conditions present in Eritrea make the malaria transmission and distribution pattern seasonal and irregular commonly characterized by predominant focal widespread epidemics. In Eritrea, malaria was recognized as endemic disease first and foremost during the Italian and British colonial period from the mid-1920s to the late 1950s. About 3.6 million (71%) of Eritrean population live in areas conducive for high transmission of malaria in which greater than one case is recorded for every 1000 people [5]. A 2017 WHO report on Eritrea indicate the most common parasite species in Eritrea is P. falciparum (70%) and major Anopheles species is An. Arabiensis which is also known to be endophilic [5]. Eritrea is inhabited by more than 13 different species of Anopheline mosquitoes all capable of spreading the disease and with varying geophysical habitats [6]. Also, inoculation rates have a high seasonal variability, with peak inoculation rates during the rainy season and minimal or no transmission during the dry season [7].

In last decades, several devastating outbreaks of malaria has been recorded in horn of Africa especially in Eritrea and Ethiopia. The 1958 malaria outbreak was one of the most memorable epidemics which resulted in an estimated of 150,000 deaths and three million cases [8, 9]. Until recently, malaria was one of the top five leading causes of mortality and morbidity in Eritrea [10].

Previously, malaria was recognized to have key impact on the socio-economic development of Eritrea. It affects the productive labor force tremendously especially during outbreaks as about 7 to 13 days are lost per episode of malaria [11]. The average cost for managing and treating an episode of uncomplicated malaria using the new effective drugs artemisinin combination therapy(ACT) is about US$2.40 and about US$7.00 for severe cases [12]. These medical intervention costs are significantly beyond the capacity of a country with a per capita GDP below US$ 200.

In the previous year of 2017, almost 10 million US dollars financed from various sources including the Eritrean Government, World Bank, WHO, UNICEF, USAID and other Global Funds have been expended in Eritrea for different diagnostic tools and intervention policies and strategy plans for malaria [5]. Though slide positivity rate remain somehow constant at levels below 25%, RDT positivity rate which was present at 75% declined substantially to low levels [5].

The key components of the malaria control program in Eritrea include diagnosis and treatment of cases, early detection of epidemics through appropriate surveillance and application of selective vector control measures and strengthening the information system to facilitate the prevention [13]. Patients admission and laboratory registers in Hospital are important sources of malaria data because they are readily accessible and can provide valuable indicators on the current situation of malaria at a lower cost. Those materials are beneficial not only for epidemiological surveillance but also for planning malaria control programs and assessing the impact of health services. If properly utilized, this information extracted from these data records will advocate the decision makers involved in malaria control to intensify control interventions timely and effectively [14].

However, currently information regarding gender, age and locality based prevalence of malaria is inadequate in Eritrea despite of the identification of the disease burden since the early of last century. Therefore, the purpose of the present study was to determine the prevalence of malaria from peripheral blood smear examination from the Nakfa Hospital found in Nakfa town. The present study outcome and implication could serve as valuable resource for malaria control and surveillance strategy in the upcoming years.

## Methods

### Study design and setting

The study was conducted at Nakfa Hospital located in Nakfa town found between Latitude 16.6655 & Longitude 38.4768, North Eritrea, one of malaria endemic area of the country. The altitude of Nakfa town ranges from 2100 to 2400 m above sea level. This study included patients who visited the hospital from September 2016 to August 2019. Based on figures released by public health campaign programs in 2019, the Nakfa subzone comprises approximately 50,335 people living within its 10 administrative units. Most of inhabitants have a semi-nomadic way of life and are involved in grazing animals. Five years average annual rainfall for that place was 378 mm with unimodal rainy season. The monthly average maximum and minimum temperatures were 28°C & 12°C for five consecutive years (2015-2019). Relative humidity data for the town is not available. Nakfa subzone includes one hospital and 10 health stations distributed over each administrative unit and health representative in each village. However, laboratory based medical treatment services are accessible at the hospital only. Nakfa hospital has 5 laboratory technicians to perform diagnosis of malaria.

A total of 5826 blood films were prepared from clinically suspected malaria cases by collecting 3 ml of blood using venipuncture technique in full aseptic conditions. Peripheral thick and thin blood smears were prepared on same slide and stained with 10% Giemsa solution. Stained slides were considered positive if at least one malarial parasite was detected, and negative when no parasite is present in 200 high power fields. Also the thin smear was used to identify the species of pre-detected parasites which was examined independently by two lab technicians and was also cross referenced with rapid diagnostic tests. The data for the current study was extracted using laboratory registers that included all malarial cases admitted to Nakfa hospital between September 2016 and August 2019. All demographic details of subjects were also retrieved from laboratory records.

### Statistical analysis

The data was analyzed statistically using SPSS package version 20. Descriptive statistics was used to evaluate the data and results were displayed in terms of tables, graphs and percentages. A 95% of confidence interval & p-value was applied as a level of significance.

## Results

During the study period, a total of 5826 blood smear were examined to detect presence of malarial parasite. The total number of males and females was 3139 (53.8%) and 2687 (46.2%), respectively. The overall prevalence of smear positive malaria cases was 33.0% (1921 cases out of 5826). Among patients who underwent diagnostic testing for malaria, males 58.19% (1118/1921 cases) were more prone to have a positive malaria smear than females 41.8% (803/1921 cases). The male to female ratio was found to be 1.39:1. A statistically significant association between number of positive blood films and male patients was observed in Chi-square analysis (p-value = 0.000) (Table 1).

**Table 1:**
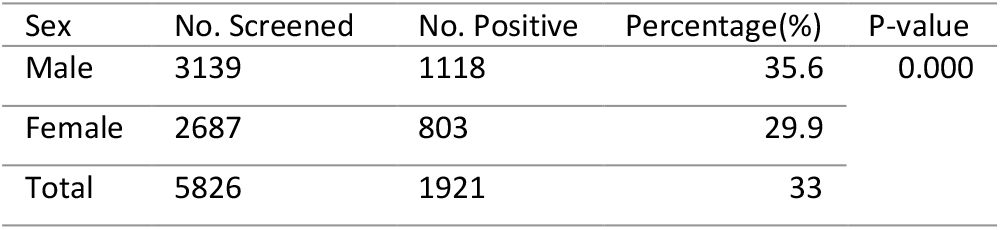
The overall Blood film positive rate of malaria in relation to sex at Nakfa Hospital, Eritrea, 2016-2019

| Sex | No. Screened | No. Positive | Percentage(%) | P-value |
| --- | --- | --- | --- | --- |
| Male | 3139 | 1118 | 35.6 | 0.000 |
| Female | 2687 | 803 | 29.9 |  |
| Total | 5826 | 1921 | 33 |  |

All confirmed malaria cases were categorized into three age groups as <5 years, 6-15 years and >15 years old (Figure 1). A higher prevalence of malaria was observed in the adult age group [(35%)1127/3231 cases] compared with children under 5 years old children [(27%)339/1240 cases]. In chi square analysis, age was also significantly associated with acquisition of malaria (p-value= 0.000) (Table 2).

**Table 2:** Blood film positive rate of malaria by age groups in patients who visited Nakfa Hospital, Eritrea, 2016-2019

| Age | No. Screened | No. Positive | Percentage(%) | P-value |
| --- | --- | --- | --- | --- |
| <5 | 1240 | 339 | 27.3 | 0.001 |
| 6-15 | 1355 | 455 | 33.6 |  |
| >15 | 3231 | 1127 | 34.9 |  |
| Total | 5826 | 1921 | 33.0 |  |

**Fig. 1.**
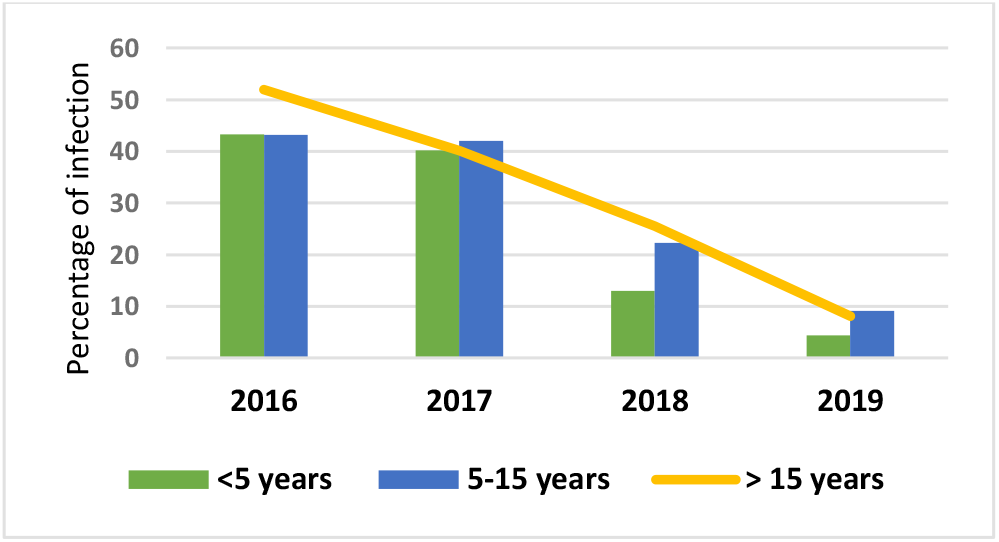
The distribution of malaria cases in different age groups and infection in Nakfa Hospital from 2016 to 2019

The highest prevalence of malaria cases was found in the year 2016 in which 489 patients were positive out of 1003 visitors corresponding to 48.8%. A comparative but lower incidence was also observed during the next year of 2017 with rate of 41% (1087 cases out of 2680). However, the year 2019 had the least number of cases where only 61 cases were detected. This difference in prevalence was statistically significant in chi square analysis (p-value=0.000) (Table 3).

**Table 3:** Blood film positive rate of malaria in Nakfa Hospital, Eritrea from 2016-2019

| Year | No. screened | No. Positive | Percentage(%) | P-value |
| --- | --- | --- | --- | --- |
| 2016 | 1003 | 489 | 48.75 | 0.000 |
| 2017 | 2680 | 1087 | 40.89 |  |
| 2018 | 1330 | 284 | 21.35 |  |
| 2019 | 813 | 61 | 7.50 |  |
| Total | 5826 | 1921 | 33.0 |  |

Infections of *P. vivax, P. falciparum* alone and mixed infection were detected in blood smears (figure 2). *P. vivax* had the highest prevalence [79.12% (1520/1921 cases)] compared to that of *P. falciparum* [21.26% (414/1921)]. During the study period, no case of *Plasmodium malariae* or *Plasmodium ovale* infection was observed. However, 13 cases of mixed infection (*P. vivax* and *P. falciparum*) were detected which contributed to 0.67% of infection (Table 4).

**Table 4:**
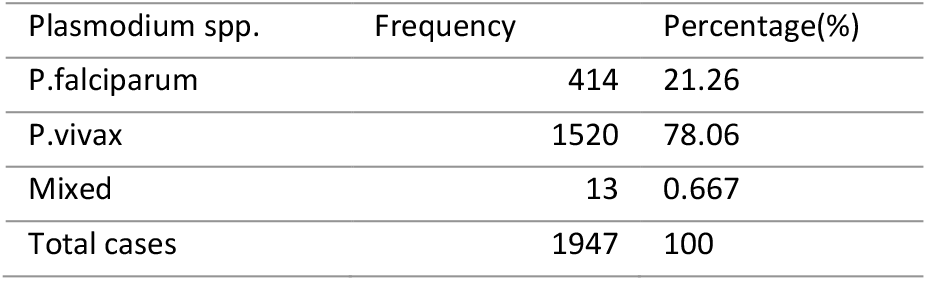
Prevalence of plasmodium species in Nakfa Hospital, Eritrea, 2016-2019

| Plasmodium spp. | Frequency | Percentage(%) |
| --- | --- | --- |
| *P.falciparum* | 414 | 21.26 |
| *P.vivax* | 1520 | 78.06 |
| Mixed | 13 | 0.667 |
| Total cases | 1947 | 100 |

**Fig. 2:**
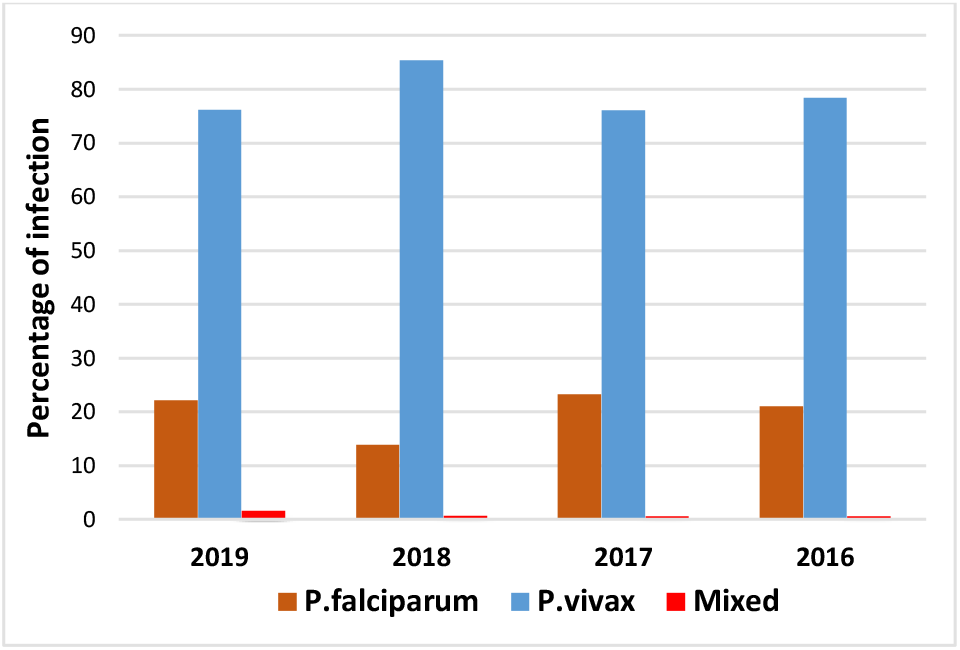
Plasmodium species trend in Nakfa Hospital from 2016 to 2019

Moreover, malaria was also statistically associated with seasonal variation (p-value<0.005). The highest prevalence was noted during Summer (40%) and Autumn (39.52%) whereas Spring had the lowest frequency of cases (143 cases out of 1000[14.3%]). Similarly, months of July (45%) and September (40.6%) had been noted to have the maximum number of cases. But months of February (12.36%) and March (12.2%) were observed to have the lowest prevalence (Table 5).

**Table 5:**
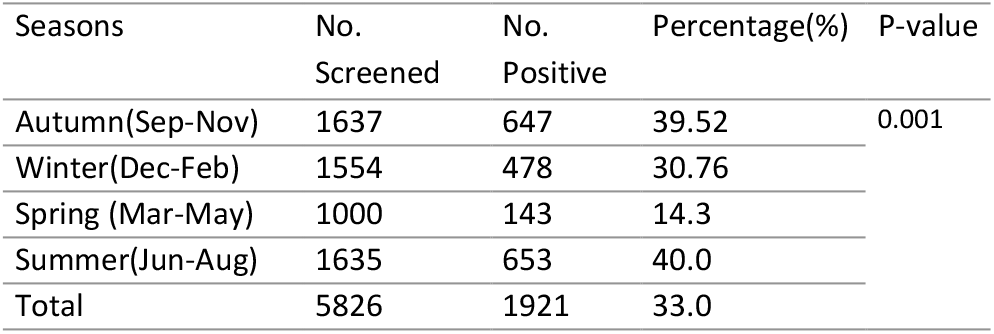
Blood film positive rate of malaria by age groups in patients who visited Nakfa Hospital, Eritrea, 2016-2019

| Seasons | No. Screened | No. Positive | Percentage(%) | P-value |
| --- | --- | --- | --- | --- |
| Autumn(Sep-Nov) | 1637 | 647 | 39.52 | 0.001 |
| Winter(Dec-Feb) | 1554 | 478 | 30.76 |  |
| Spring (Mar-May) | 1000 | 143 | 14.3 |  |
| Summer(Jun-Aug) | 1635 | 653 | 40.0 |  |
| Total | 5826 | 1921 | 33.0 |  |

Locality based analysis show around 70% of cases were from patients living in and at the peripheries of Nakfa town. However, marked prevalence were also observed in rural villages like Meo which had 17.78 % of total cases (Figure 3).

**Fig. 3.**
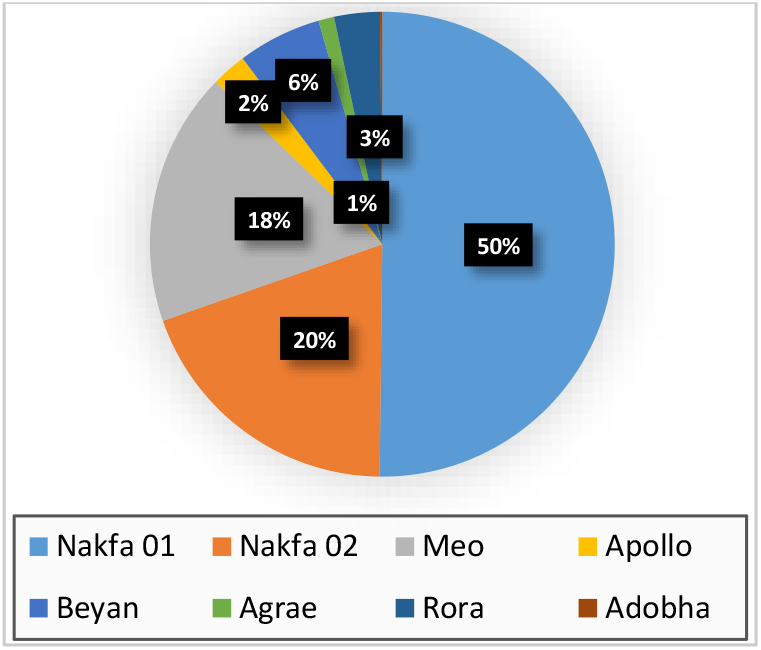
Locality based distribution of malaria in Nakfa Hospital from 2016 to 2019

## Discussions

Malaria is a common parasitic infection affecting huge populations residing in endemic areas and continues to remain a serious public health problem in sub-Saharan Africa. Malarial parasites account for high morbidity and mortality in children and pregnant women present in tropical countries. The overall prevalence of malaria in this study was 33.0% where a total 1921 confirmed malaria cases were detected in nearly three years’ retrospective study. Comparatively, the Nakfa area is regarded as one of malaria endemic areas of the country as indicated in WHO Eritrea profile [5]. Due to absence of similar published articles, though result cannot be compared to other counterpart Eritrean areas, the prevalence was higher in contrast to studies conducted in south Ethiopia [15, 16, 17] but lower compared to other parts of Ethiopia [18]. These variations could be due to altitude differences and climate diversity which directly relate to reproduction of Anopheles species.

Among patients who had diagnostic test for malaria, males [35.6% (1118/3139 cases)] were more at risk to have positive blood film than females [29.9% (803/2687 cases)]. The chi-square test analysis clearly showed that there was a significant association between male patients and acquisition of malaria (p-value= <0.05). For reasons related to social way of living in this region, males are usually out of the city for education, employment including grazing of animals and farming and other activities which leave the male population more active during evening time for their day to day outdoor activity [19, 20, 21]. This was also in agreement with other studies conducted in Ethiopia [22].

The least affected age group was children (< 5 years) (27.3%) followed by the age group 6-15 which had prevalence of 33.6%. The age group of subjects greater than 15 years old had the majority of cases (34.9%) compared to its contraries. This result was in consistence with study conducted in different part of the world [19, 23]. This can be attributed to greater portion of subjects diagnosed being in the age above 15 years which are also commonly breadwinners of their families spending most of their time especially evenings outdoors when the peak biting activity of Anopheles is seen [24].

In this study, *Plasmodium vivax* constituted the most predominant malaria infections [78.06% (1520/1921cases)], while markedly lower *Plasmodium falciparum* was also present [21.26 % (414/1921 cases)]. This is however inconsistent with the overall WHO report for Eritrea that reveal about 70% of malaria infections are related to *p. falciparum*. But the same report also refers Nakfa area as one of the leading risk regions for p. vivax caused malaria in the country [5]. This pattern of infection can be explained by how temperature affects the life cycle of the malaria parasite. The time required for the parasite to complete its development in the gut of the mosquito is about 10 days, but it can be shorter or longer than that depending on the temperature. The time needed for the parasite to complete its development in the mosquito, decreases to less than 10 days as temperature increases from 21°C to 27°C, with 27°C being the optimum. The maximum temperature for parasite development is 40°C. In areas like Nakfa whose average temperature below 18°C, the life cycle of *P. falciparum* in the mosquito body is limited. The minimum temperatures are between 14–19°C, with P. vivax surviving at lower temperatures than P. falciparum which correspond to high p. vivax infection rate in Nakfa area [25]. However, it is essential to note that the predominant malaria in Africa is related to p. falciparum accounting for almost 100% of malaria disease in most of sub-Saharan region. Furthermore, 0.66% of mixed malarial infection of both *P. vivax* and *P. falciparum* were also detected. This finding is also in agreement with other study in Ethiopia [15] and India [26].

A very virulent epidemic was observed in summer 2017 in the months of July and August which increased the rate of decrease in transmission of infection compared to the previous year. Out of the total 2680 visiting patients, 1087 had a positive malaria blood film test corresponding to 41% of all patients. The outbreak was controlled in few months through early diagnosis and treatment protocols followed at health facility and community levels and proper management of severe malaria at the higher health services. To reduce man-mosquito contact, insecticide-treated nets were distributed at every administrative unit in Nakfa area. Also programs that increase community awareness were organized through the promotion of information, education, and communication.

Moreover, seasonal variations of malaria prevalence have been observed in which maximum number of cases was seen in the month of September [227 cases out of 559 (40.6%)] and minimum cases [only 51 cases out of 418 (12.2%)] were detected in month of March. This is related to the three main climatic factors that directly affect malaria transmission, i.e. temperature, rainfall and relative humidity which are variable at different times of year [25]. This trend of seasonal variation was found to be similar to studies conducted in different parts of the world [27].

Almost around 70% of cases were reported from patients living in and at the peripheries of Nakfa town. This may be attributed to the biggest portion of hospital visiting subjects being from the town itself. However, marked prevalence were also observed in rural villages like Meo which had 17.78% of total cases (Figure 3).

## Conclusions

Eritrea is one of the few countries which has made a profound progress on the three programs of communicable diseases control program namely HIV, TB and Malaria, which emphasizes the value placed on ensuring health for all populations. Moreover, marked reduction trends were observed in morbidity and mortality throughout the past years. However, vigilant surveillance is required especially during malaria transmission peaks from July to October which also overlap with harvesting seasons in Eritrea. Therefore, health planners and decision makers need to organize intensive health education to increase community awareness via promotion of information and communication.

## Data Availability

The corresponding author confirms that all data underlying the findings
are fully available upon request. All relevant data are within the
manuscript.

## Abbreviations

WHO: world health organization;
p. falciparum: plasmodium falciparum;
p.vivax: plasmodium vivax

## Ethical approval and consent to participate

The data was collected after ethical clearance was obtained from the Department of Clinical Laboratory Science, Asmara College of Health Sciences and Ministry of Health, State of Eritrea. After discussing the objective and methods of the study, a written permission was obtained from Nakfa Hospital before the data collection period. In order to maintain anonymity and ethical confidentiality, all patients were given codes during the data extraction process and patient names were not extracted from the register.

## Consent for publication

Not applicable.

## Availability of data and materials

The corresponding author confirms that all data underlying the findings are fully available upon request. All relevant data are within the manuscript.

## Authors’ Contribution

Y. K participated in conception and design of the study, data collection and analysis, interpretation of the findings, and wrote the manuscript. SM participated in conception and design of the study, data collection and analysis. MT participated in data collection and analysis, interpretation of the findings and revised the manuscript. All authors approved the final manuscript.

## Acknowledgements

The authors wish to thank all staff members of Nakfa Hospital, North Eritrea for their timely consistent help and support who assisted in retrieving the case notes to carry out this research work.

